# Conceptualization, operationalization, and utilization of race and ethnicity in major medical journals 1995-2018: a systematic review

**DOI:** 10.1101/2022.03.07.22271661

**Authors:** Rae Anne M. Martinez, Rachel E. Wilbur, Nafeesa Andrabi, Andrea N. Goodwin, Natalie R. Smith, Paul N. Zivich

## Abstract

**Background:** Systemic racial and ethnic inequities continue to be perpetuated through scientific methodology and communication norms despite efforts by medical institutions.

**Purpose:** To characterize methodological practices regarding race and ethnicity in U.S. research published in leading medical journals.

**Data source:** Articles published in Annals of Internal Medicine, BMJ, JAMA, The Lancet, and NEJM from 1995-2018 were sampled via PubMed.

**Study Selection:** All original, human subjects research conducted in the U.S.

**Data Extraction:** Information on definition, measurement, coding, use in analyses, and justifications was collected.

**Data Synthesis:** The proportion of U.S. medical research studies including race and/or ethnicity data increased between 1995 and 2018. No studies defined race or ethnicity. and most did not state how race and/or ethnicity was measured. Common coding schemes included: “Black, other, White,” “Hispanic, Non-Hispanic,” and “Black, Hispanic, other, White.” Race and/or ethnicity was most often used as a control variable, descriptive covariate, or matching criteria. Under 30% of studies included justification for their methodological choices regarding race and/or ethnicity.

**Conclusions:** Despite regular efforts by medical journals to implement new policies around race and ethnicity in medical research, pertinent methodological information was systematically absent from the majority of reviewed literature. This stymies critical disciplinary reflection and progress towards equitable practice.

## INTRODUCTION

Following global protests for racial equity, an increasing number of health researchers are studying racism as a fundamental cause of morbidity and mortality. Such investment is long overdue. However, racism-focused work must be coupled with sound methodological practices regarding the social constructs of race and ethnicity. Effectively using these constructs is integral to documenting and understanding how systems of racism and ethnocentrism affect health. Unfortunately, practices surrounding race and ethnicity in medical research are often deficient regarding definitions, measurement, coding, analysis, and interpretation of findings. Perpetuating problematic methodological practices maintains an ethnocentric status quo and may contribute to challenges in understanding how racism affects health, ultimately hindering effective and equitable healthcare and policy-making.

Debates over appropriate methodological approaches to race and ethnicity in health are longstanding. In the 1990s, researchers challenged many methodological decisions, including the necessity of racial and/or ethnic data, construct definitions, measurement choices, appropriateness of coding schemes, and role of variables in analyses (1-8). At the time, Thomas LaVeist (1996) argued that racial and ethnic data retained high utility for health research. He challenged health researchers to “do a better job” of conceptualizing race, understanding nuances of racial and ethnic measurements, and interpreting findings with care in order to help reduce health disparities in the U.S. (9). Recent work in surgery and oncology has identified infrequent reporting of race and ethnicity data (10-12), however, no comprehensive systematic review of the state of these methodological practices in medicine over time currently exists.

The present study seeks to fill this gap by systematically reviewing trends in methodological practices regarding the conceptualization, operationalization, and utilization of race and ethnicity in U.S. medical literature. By examining publications in influential medical journals over the past quarter-century, we document the state of medicine’s methodological norms and identify patterns of disciplinary practices that may reify misconceptions about race and ethnicity, with implications for scientific quality, reproducibility, and equity. In total, we investigated five core questions using a sample of U.S. medical publications: 1) What proportion of studies incorporate data on race and ethnicity? 2) What proportion provides conceptualization of race and ethnicity? 3) How is race and ethnicity data operationalized? 4) How is race and ethnicity data utilized in analyses? And 5) Do the authors justify their methodological decisions regarding race and ethnicity in publication?

## METHODS

This study is a methodological systematic review under Munn et al.’s taxonomy (13), as the foundational methodological treatment (i.e., definitions, measurement, coding, analytical use, and scientific justifications) of two key variables - race and ethnicity - is the focus of this investigation. We define race as a social and political construct whereby social meanings (e.g., beliefs about ability, health, worth, etc.) are assigned to arbitrary phenotypes and which capture differential access to power, opportunities, and resources in a race-conscious society (14, 15). Similarly, we define ethnicity as a social construct, stemming from a sense of belonging over shared cultural elements (e.g., language, religion, traditions, values) and/or of place (e.g., national origin) (14, 16). Both race and ethnicity are contextually, temporally, and geographically specific; neither race or ethnicity are determined by biology (17-19). For the purpose of this review, “Hispanic” and “Latino/a/x/e” are defined as a pan-ethnic identities, not as racial identities. Furthermore, “African American” is defined as an ethnic identity and is not synonymous with “Black.” See Appendix 1 for additional background and rationale. Capitalization practices were not collected from sampled articles; however, we follow the AMA capitalization style guidelines and capitalize all racial and/or ethnic terms in this article (20).

### Data sources and searches

The target articles under study include all U.S.-based, original, human subjects medical research published in *Annals of Internal Medicine, BMJ, Journal of the American Medical Association* (JAMA), *The Lancet*, and the *New England Journal of Medicine* (NEJM) between Jan 1, 1995 and Dec 31, 2018 (Figure 1). Journals were selected based on impact factor and reputation, consistent with other methodological systematic reviews (21-23).

**Figure 1.**
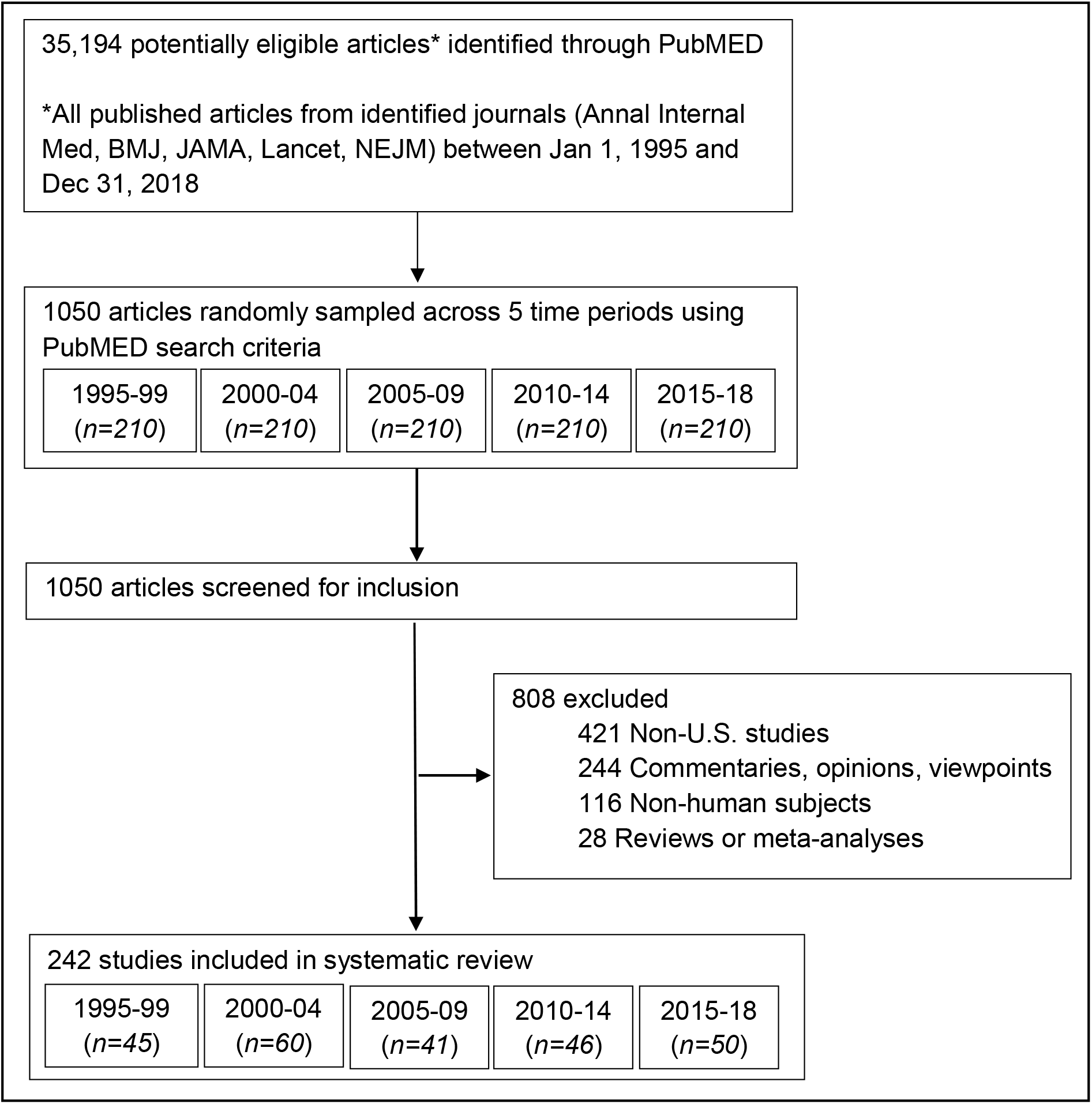
Study selection. Total population of articles includes all articles published in the five identified journals between Jan 1 1995 and Dec 31 2018. In total, 35194 articles were returned; this includes articles that do not meet study eligibility criteria (i.e., US-based, original human subjects research).

Studies were identified by searching PubMed for empirical work published between Jan 1, 1995 and Dec 31, 2018. To reduce ineligible articles the following search terms were used: (English[Language]) NOT (Letter[Publication Type]) NOT (Comment[Publication Type]) NOT (Editorial[Publication Type]) NOT (Review[Publication Type]) NOT (News[Publication Type]) NOT (Case Reports[Publication Type]) AND ((“United States”[MeSH]) OR (“United States”[tw]) OR America[tw] OR “U.S.”[tw] OR “US”[tw]). Given the number of articles returned by the original search (35,194; Figure 1) and the richness of the data we aimed to collect, we took a stratified random sample of 210 articles from five, five-year periods (1995-1999; 2000-2004; 2005-2009; 2010-2014; 2015-2019; 1050 articles total). Data collection occurred between July 2019 and November 2021.

### Study selection

All human-subjects research conducted exclusively in the U.S. was included. Non-U.S.-based research or multi-national research was excluded because of the unique social and geopolitical structures through which race and ethnicity function. We encourage researchers in other countries to conduct similar reviews using language and racial and/or ethnic categories that are important and specific to their context. Letters to the editor, commentaries, meta-analyses, and simulation studies were excluded. No restrictions were made on study outcome, exposure, or study design.

### Data extraction and quality assessment

Full details on the protocol have been reported elsewhere (23). In brief, all included articles were independently reviewed in-full by two reviewers; data were abstracted into a standardized REDCap form (24, 25). Abstraction was conducted using an existing protocol and all reviewers were primed using practice articles. Any abstraction discrepancies were discussed between the pair of reviewers, and if consensus could not be reached, were reviewed collectively by the author team. A third data quality check was conducted by the primary author. See Appendix 2 for details.

### Software

Articles were sampled with Python 3.5.2 (26) using Biopython (27) and NumPy (28) libraries. Analyses were performed in R, version 4.0.2 (29) with packages tableone (30), tidytext (31), and tidyverse (32).

### Funding

Financial support was provided in part by training grants from the Eunice Kennedy Shriver National Institute of Child Health and Human Development [T32-HD091058], National Cancer Institute [T32-CA057711], and the National Institute of Allergy and Infectious Disease [T32-AI007001] with general support from the Carolina Population Center [P2C-HD050924, P30-AG066615]. Additional pilot funding was provided by the Department of Sociology, University of North Carolina at Chapel Hill. Funding sources had no role in data collection, analysis, interpretation, or any other aspect pertinent to the study.

## RESULTS

Of 1050 screened articles, 242 were included (Figure 1). The majority of excluded articles were either international studies or commentaries (Figure 1).

Across time periods, the majority of studies were either cohort studies (range 56-73%) or randomized control trials (range 18-41%; Table 1). Most studies examined a physical or mental health outcome (range 70-80%). “Other” outcomes were the second most prevalent (16-23%) and included studies on topics such as medical training, medical errors and the prevention of adverse events, or physician decision making.

**Table 1.** Characteristics of included articles (*N=242*)

|  | 1995-99 | 2000-04 | 2005-09 | 2010-14 | 2015-18 |
| --- | --- | --- | --- | --- | --- |
| **No. Included** | 45 | 60 | 41 | 46 | 50 |
| **Study Design, No. (%)** |  |  |  |  |  |
| RCT | 9 (20) | 11 (18) | 17 (41) | 14 (30) | 14 (28) |
| Cohort | 33 (73) | 44 (73) | 23 (56) | 32 (70) | 32 (64) |
| Case-control | 2 (4) | 3 (5) | 1 (2) | 0 (0) | 2 (4) |
| Ecological | 1 (2) | 2 (3) | 0 (0) | 0 (0) | 2 (4) |
| **Study Outcome, No. (%)<sup>\*</sup>** |  |  |  |  |  |
| Health behavior | 2 (4) | 2 (3) | 1 (2) | 0 (0) | 2 (4) |
| Physical or mental | 33 (73) | 45 (75) | 33 (80) | 35 (76) | 35 (70) |
| Healthcare access | 3 (7) | 6 (10) | 3 (7) | 5 (11) | 5 (10) |
| Other | 7 (16) | 14 (23) | 8 (20) | 8 (17) | 10 (20) |
| **Sample size, No. (%)<sup>†</sup>** |  |  |  |  |  |
| <1000 | 26 (54) | 28 (42) | 13 (26) | 15 (29) | 15 (30) |
| 1000-5000 | 12 (25) | 11 (17) | 13 (26) | 9 (17) | 8 (16) |
| 5001-10,000 | 2 (4) | 4 (6) | 2 (4) | 2 (4) | 4 (8) |
| 10,001-100,000 | 3 (6) | 10 (15) | 10 (20) | 15 (29) | 9 (18) |
| >100,000 | 2 (4) | 9 (14) | 12 (24) | 7 (14) | 12 (24) |
| Missing | 3 (6) | 4 (6) | 0 (0) | 4 (8) | 2 (4) |
| **Inclusion of data, No. (%)** |  |  |  |  |  |
| Racial data | 20 (44%) | 31 (52%) | 27 (66%) | 33 (72%) | 37 (74%) |
| Ethnic data | 9 (20%) | 16 (27%) | 19 (46%) | 25 (54%) | 29 (58%) |
| **Collapsing of racial & ethnic data, No. (%)<sup>‡</sup>** |  |  |  |  |  |
| Collapsed into single construct | 9 (100%) | 13 (81%) | 18 (100%) | 21 (84%) | 25 (86%) |
| Not collapsed | 0 (0%) | 3 (19%) | 0 (0%) | 4 (16%) | 4 (14%) |

<sup>\*</sup>Study outcomes were classified as health behaviors (e.g., smoking, dietary intake, physical activity, sexual behaviors), mental or physical health (e.g., obesity, high blood pressure, cancer, depression), health care access or utilization (e.g., health insurance status, number of primary care visits, quality of care), or other. Study outcomes are not mutually exclusive and may sum to more than 100%. <sup>†</sup>Some studies listed more than one analytic sample size; values may sum to more than 100%. <sup>‡</sup>Denominator: Studies had to include both racial and ethnic data. Studies that did “not collapse” kept racial and ethnic data separate throughout the entire analyses.

### Question 1: Inclusion of racial and ethnic data

The proportion of reviewed studies that included data on participants’ race increased over time (range 44-74%, Table 1). Studies that did not include participants’ racial data do not substantially differ from the overall sample with respect to study design, study outcome, or sample size (Appendix Table 2). Over the same period, the proportion of reviewed studies that included participants’ ethnicity data has similarly increased (range 20-58%, Table 1).

**Table 2.** Measures of race and ethnicity over time, 1995-2018.

|  | 1995-99 | 2000-04 | 2005-09 | 2010-14 | 2015-18 |
| --- | --- | --- | --- | --- | --- |
| **No. Included race data** | 20 | 31 | 27 | 33 | 37 |
| **Measure of race, No. (%)** |  |  |  |  |  |
| Open-ended, self-report | 0 (0) | 0 (0) | 0 (0) | 0 (0) | 1 (3) |
| Close-ended, self-report | 1 (5) | 3 (10) | 2 (7) | 2 (6) | 7 (19) |
| Observed | 0 (0) | 0 (0) | 3 (11) | 0 (0) | 0 (0) |
| Phenotype | 0 (0) | 0 (0) | 0 (0) | 0 (0) | 0 (0) |
| Reflected | 0 (0) | 0 (0) | 0 (0) | 0 (0) | 0 (0) |
| Ancestry | 0 (0) | 0 (0) | 0 (0) | 0 (0) | 0 (0) |
| Unclear/not stated | 18 (90) | 28 (90) | 16 (59) | 20 (61) | 22 (59) |
| Open vs close-ended* | 1 (5) | 0 (0) | 7 (26) | 11 (33) | 7 (19) |
| **No. Included ethnicity data** | 9 | 16 | 19 | 25 | 29 |
| **Measure of ethnicity, No. (%)** |  |  |  |  |  |
| Open-ended, self-report | 0 (0) | 0 (0) | 0 (0) | 0 (0) | 1 (3) |
| Close-ended, self-report | 0 (0) | 3 (19) | 3 (16) | 1 (4) | 7 (24) |
| Country of origin | 0 (0) | 1 (6) | 0 (0) | 0 (0) | 0 (0) |
| Observed | 0 (0) | 0 (0) | 0 (0) | 0 (0) | 0 (0) |
| Reflected | 0 (0) | 0 (0) | 0 (0) | 0 (0) | 0 (0) |
| Ancestry | 0 (0) | 0 (0) | 0 (0) | 0 (0) | 0 (0) |
| Unclear/not stated | 8 (89) | 12 (75) | 12 (63) | 17 (68) | 15 (52) |
| Open vs close-ended* | 1 (11) | 0 (0) | 5 (26) | 7 (28) | 6 (21) |

Selections of multiple measures was allowed; percents may sum to >100%. \*For racial "open vs close-ended" race was noted as self-reported by a participant, but it was unclear if the question was open- or closed-ended. Same applies to ethnic "open vs close-ended."

Racial and ethnic data were almost always included together in the same study. Across all 149 studies which included participants’ race and/or ethnicity data, only a single study included data on participants’ ethnicity without also including data on participants’ race. When ethnicity data was included in the study, it was frequently combined with race into a single ethnoracial construct (range 81-100%, Table 1). Only 11 studies across all strata included both race and ethnicity data and kept them as separate entities.

### Question 2: Conceptualization of race and ethnicity

Across all 149 studies which included data on participants’ race and/or ethnicity, none provided a definition of either construct.

### Question 3: Operationalization

In 59-90% of articles across strata, the measurement of race was “not stated or unclear” (Table 2). In articles that indicated using “self-reported” race, it was frequently ambiguous if the measure was open-ended (i.e., free response) or close-ended (i.e., selection from preset options). Ambiguity between “open” and “closed” measures was more common in later strata (2005-09, 2010-14, 2015-18, Table 2). Use of other measures (e.g., observed, reflected, or phenotype) was infrequent or absent (Table 2).

Results for ethnicity are similar; across all strata, articles commonly lacked any information on measurement of ethnicity (range 52-89%, Table 2). Ambiguity between open and closed measures was more common in later strata (2005-09, 2010-14, 2015-18, Table 2), and other measures (e.g., country of origin) were rare.

Coding schemes were collapsed across sampling strata and examined by use of a strictly racial, ethnic, or a collapsed ethnoracial construct. Racial and ethnoracial coding schemes were more heterogeneous, while ethnic coding schemes were more similar (Table 3). Although “non-White, White” and “nonWhite, White” are functionally the same, we made no attempt to collapse coding schemes based on similarity due to concern about the subjectivity of those decisions. The most common racial coding schemes reflected predominantly binary racial framing centering “Whiteness,” while ethnic coding schemes primarily centered on “Hispanic” or “Latino” binary coding. In the most common ethnoracial coding schemes “Hispanic” - an ethnic group - is compared to the racial categories of “White” and “Black.” Ethnic, racial, and ethnoracial codings all included “ns (not stated),” where no information was provided in the article about how participants’ racial and/or ethnicity data was re-coded for the study. Appendix Tables 3 and 4 contain the complete list of racial and ethnoracial coding schemes, respectively.

**Table 3.** Most frequent coding schemes.

|  | No. | (%) |
| --- | --- | --- |
| **Racial coding schemes** |  |  |
| Black, other, White | 9 | (15) |
| Black, White | 9 | (15) |
| non-White, White | 6 | (10) |
| nonWhite, White | 5 | (8) |
| ns (not stated) | 5 | (8) |
| White | 5 | (8) |
| Asian, Black, other, White | 4 | (6) |
| Black | 3 | (5) |
| **Ethnic coding schemes** |  |  |
| Hispanic, non-Hispanic | 5 | (42) |
| ns (not stated) | 4 | (33) |
| Hispanic | 1 | (8) |
| Hispanic or Latino, not Hispanic or Latino | 1 | (8) |
| Hispanic origin | 1 | (8) |
| **Ethnoracial coding schemes** |  |  |
| Black, Hispanic, other, White | 9 | (14) |
| Asian, Black, Hispanic, other, White | 5 | (8) |
| Black, Hispanic, White | 4 | (6) |
| Black, other, White* | 3 | (5) |

### Question 4: Use in analyses

Race and ethnicity were predominantly classified as “not of interest” in analyses (i.e., used as a descriptive covariate, confounder, or matching criteria; range 64-84%; Appendix Table 5). Only four studies across stratum used race and/or ethnicity as an exclusion criterion, two of which restricted analysis to solely White participants. In 10-25% of studies across stratum, race and/or ethnicity were “of interest” (e.g., specific group comparisons, effect measure modification, or predictive variable).

### Question 5: Justification

Approximately 30% of the 149 studies across strata which included participants’ racial and/or ethnic data provided a justification for at least one of their decisions surrounding race and/or ethnicity (e.g., the relevance of race and/or ethnicity to the study question, choice of measure, generation of coding scheme, and why an analytical approach or use of the variable was appropriate; data not shown). No studies provided justifications for the selection of a particular measure (e.g., selection of close-ended, self-report question over an open-ended, self-report question). Three studies referenced National Institutes of Health or other institutional guidelines with respect to decisions making on measurement and coding. As in Castro *et al*. (2014), authors explained “race was assessed by participant self-report, using National Institutes of Health race/ethnicity reporting standards and categories” (p.2085-2086) (33).

## DISCUSSION

We systematically review methodological practices regarding the conceptualization, operationalization, and use of race and ethnicity in U.S. medical research published in prominent journals between 1995-2018. We found that information specific to race and ethnicity was routinely, if not systematically, absent from articles. While inclusion of racial and ethnic data has increased since 1995, no studies defined either construct and most did not describe how race and/or ethnicity was measured. Occasionally, the coding schemes of racial and ethnic variables were even omitted. Most studies across time periods did not provide scientific justification for their choices with respect to race and/or ethnicity.

Scientific rigor relies on replication and validation, which is rendered impossible if core methodological decisions are not clearly communicated. Core methodology includes information on definitions, measurement, and coding of variables, as well as scientific rationale. Absence of such information may impact interpretation of findings or their translation into interventions, especially when it is unclear who is under study and why. Lack of basic information on methodology threatens our ability to conduct responsible and rigorous science.

### Scientific and cultural racism

Journal word limits provide a potential structural explanation for lack of clarity regarding race and ethnicity. Descriptions of methodological choices regarding race and ethnicity may compete with information on foundational literature, study design, exposure, outcome, results, or interpretations for inclusion.

The absence of information could also reflect a misguided belief in the presumed universality of race: that what race is and is not, the number of racial groups, boundaries between racial groups, and the “scientific relevance” of race to medical research are invariably understood. If race and ethnicity are universally understood across temporal, socio-cultural, and geopolitical contexts, then “race” does not need explanation or justification. Race, however, is not universal. Rather, what “race” is, the number of and boundaries between “racial groups,” and mechanisms by which the multilevel system of racism operates are deeply contextual. A large body of literature has theorized on how the social construction of racial and ethnic categories is historically situated and changes over time and place (14, 34-38).

The U.S., for example, is a nation explicitly designed to prioritize the life chances of a single group of people. As a settler-colonial state which achieved global financial power through slave labor and imperialism, the structures which continue to support the political, financial, judiciary, and educational systems maintain a hierarchical status quo based on established racial groups (39). Racism may be globally pervasive, but the structure of the system and the experience of living within it is different in the U.S. than it is in Mexico, Brazil, South Africa, India, or any other country.

Combatting scientific racism in medicine, in part, requires naming the methodological assumptions behind the treatment of race and/or ethnicity in medical research. For over 150 years, medicine as a discipline actively reified the biological essentialist definition of race - that perceived behavioral and health differences between “racial groups” were true, immutable, and inherent to an individual’s genetic makeup (40). By routinely justifying biological essentialism with pseudoscientific evidence, race became “common sense” and perceived as part of the natural world (40). This idea has so deeply infiltrated scientific institutions and thought, that it remains present today despite the scientific process demonstrating the falsity of these claims. Within these structures, medical research in the U.S. has historically adhered to practices which harm subordinated groups as research subjects while the knowledge produced by these research practices most benefits those of the dominant group (41, 42). This practice contributed to the current state, in which racial and ethnic minorities are often systematically excluded in medicine, as both research participants and researchers (43-47). Thus, medical knowledge is predicated on only some bodies, cultures, and experiences (48, 49). The lack of diverse perspectives contributes to the perpetuation of unconscious bias and racist practices in medicine.

### Institutions and structure

Over the years, journals and other institutions have developed communication guidelines around race and ethnicity. The International Committee of Medical Journal Editors (ICMJE) developed two such recommendations in 2004, namely that 1) the inclusion of racial and ethnic data is explicitly motivated and 2) the measurement of race and ethnicity is clearly explained (50, 51).

All of the journals sampled in our study aim to follow the standards set forth by ICMJE (52). However, for U.S.-based human subjects research published in these journals, adherence appears limited. After 2004, most studies still did not include information on how race and/or ethnicity was measured. Even considering the possibility of a lag between the release of new standards and the publishing of articles following those standards, adherence is low. Furthermore, few articles included justifications for decisions surrounding race and ethnicity.

Editors and specific medical journals have further echoed and elaborated upon the ICMJE recommendations. Following the 2004 update, former JAMA Deputy Editor Dr. Margaret Winker introduced expanded ICMJE guidance specifically for network journals, calling for authors to provide details on (1) who assessed an individual’s race, (2) whether self-designation options were “open” or “closed,” (3) what the closed self-designation categories were, (4) if and how closed self-designation categories were combined, and (5) the rationale or relevance of race and ethnicity to a particular study (53). In supplemental analyses, there is minimal evidence of adherence to these additional higher standards among sampled JAMA articles (Appendix 3). Recently, the AMA has released more explicit policies (54, 55).

### Actions for improvement

Previous work in medicine and adjacent disciplines has provided suggestions for improvement (23, 56-59). We build on this work by calling for clear communication of these improved practices, including definitions, measurement, coding, use, and justifications. This is not a radical position. We simply argue that race and ethnicity should be given the same interrogation and justification as other variables, and that this be clearly communicated in publication.

We urge health researchers to follow existing guidelines and implore medical journals and editors to implement mechanisms for accountability to these standards. For example, authors could be prompted to certify at submission that they have adhered to ICMJE or AMA guidelines. At the peer-review level, additional training could be implemented to ensure that reviewers are confident in recognizing whether a manuscript meets criteria. We further encourage out of the box thinking to overcome structures; for example, pertinent details on race and ethnicity could be without word count, similar to human subjects statements or acknowledgements. Conducting annual reviews of policy adherence across medical journals could ensure that baseline benchmarks are being met.

Responsibility for meeting disciplinary standards of research falls on both medical journals and authors, as both are ultimately in service of patients and study participants. As “key players in the production of knowledge” (p.1288) and gatekeepers of research dissemination, editors and medical journals are in a unique position to ensure adherence to stringent scientific communication norms (60). In particular, prominent medical journals, by setting and requiring adherence to guidelines on clear communication, may influence disciplinary-wide standards. For authors, meeting these standards may require critical thought and conscious decoupling from earlier norms of conducting and reporting race and ethnicity in medical research.

### Limitations

The abstraction from sampled articles is imperfect. The data retain a degree of subjectivity, despite protocols to standardize data entry and data quality checks. This is perhaps most true for the data on scientific justifications. Data abstractors were instructed to be as broad as possible when collecting information on justifications, thus data may be an overestimate of articles which included at least one justification.

Second, it is possible that recent attention to addressing racism and ethnocentrism broadly has resulted in a renewed effort to “do a better job.” Subsequently, methodological practices and the communication thereof may have substantially shifted between Jan 1, 2019 and today. Finally, we did not review supplementary materials. If information on definitions, measurement, coding, or scientific justifications was included in supplements, they were missed.

## Conclusion

Interventions aimed at addressing racism as a fundamental cause of disease in the U.S. must be based on unassailable research achieved through strict methodological rigor. Quality science enables knowledge democracy and health equity by providing a strong evidence base for changes in medical practice and policy. Dismantling systematic oppression in medicine requires clear, critical, and honest communication around the use of race and ethnicity data in medicine. Collectively, the health research community needs to hold each other accountable to continue improving how race and ethnicity are conceptualized, operationalized, and utilized in medical research. This should be one element in a holistic, multipronged approach to addressing racism and health inequity which also centers additional systems reforms.

## Supporting information

Supplemental Files (ALL)

## Data Availability

REDCap data entry form and full list of articles will be made available upon request with publication. Please contact the corresponding author for data inquiries.

## CONTRIBUTORS

RAMM conceived of the study and directed its implementation, including quality assurance and control. All authors contributed to study design and data acquisition. Authors RAMM, NRS, and PNZ conducted analyses. RAMM and REW wrote the primary draft; all other authors (NA, ANG, NRS, PNZ) contributed to further drafts and edits. All authors had full access to and verified the data.

## ACKNOWLEDGEMENTS

We are thankful to Dr. Allison E. Aiello and Dr. Robert A. Hummer for their guidance and support. We are indebted to Denise Mitchell for their assistance in data collection. NRS contributed to this work while at the University of North Carolina and is now a postdoctoral fellow in the Harvard TH Chan School of Public Health Department of Social and Behavioral Sciences.

