## Supplemental Files (ALL) for "Conceptualization, operationalization, and utilization of race and ethnicity in major medical journals 1995-2018: a systematic review"

### SUPPLEMENTAL INFORMATION

#### Table of Contents

*Appendix 1. Race and ethnicity* 2

*Appendix 2. Data extraction and analysis* 5

*Appendix Table 1. Definitions of race and ethnicity dimensions* 7

*Appendix Table 2. Characteristics of studies without race data (N=94)* 8

*Appendix Table 3. Racial coding schemes (N=62)* 9

*Appendix Table 4. Ethnoracial coding schemes (N=86)* 10

*Appendix Table 5. Role of race and/or ethnicity in analyses, 1995-2018* 12

*Appendix 3. Secondary analysis: JAMA publications* 13

**Appendix 1. Race and ethnicity**

We acknowledge that language around the constructs of race and ethnicity is constantly shifting. It is therefore integral to specify terms and trace the origins of pertinent language in order to accurately convey assumptions that are made. We align with the social constructionist paradigm which recognizes that both racial and ethnic groups are socially created rather than biologically defined realities, and that society assigns social meaning to phenotypical differences or group traditions.

Race

We define race as a social and political construct whereby social meanings (e.g., beliefs about ability, health, worth, etc.) are assigned to arbitrary phenotypes which capture differential access to power, opportunities, and resources in a race-conscious society.[1] Present-day racial categories are meaningful in that they reflect the historical, political, and social processes that shape lived experience in the United States.[2]

Ethnicity

Within the scope of this study we define ethnicity as the subjectively felt sense of belonging based on the belief in and experience of shared culture, common ancestry, language, nationality, or religion.[3,4]

Panethnicity

Panethnicity is the grouping together of different ethnicities that are similar based on geographical origin, language, and religion.[5,6]

Ethnorace

Ethnorace or ethnoracial identity is a recently proposed framework, which argues that racial characteristics (e.g., skin color, hair texture, bone structure) and ethnic characteristics (e.g., language, religion, nativity) represent distinct dimensions of social stratification that may work together in particular contexts to shape an individual's experience and identity. From a practical methodological perspective, this framework offers a theoretically sound and motivated approach to integrate ethnic and racial data into one variable or concept that confers comparable meaning across these groups. Shifting towards ethnoracial classification may also offer scholars improved ability to understand the nature, extent and consequences of today's U.S. population given increased migration, growing nonwhite US population and new ethnoracial groups.[7]

Specific Examples

African American vs Black:
We draw a conceptual distinction between African American and Black, noting that they are not synonyms. We conceptualize Black as a racial identity, while we consider

African American as a US-centered ethnicity that captures a shared heritage or history, such as ancestral legacy of forced removal and enslavement.[8,9]

Hispanic & Latino/a/x/e:

Hispanic refers to individuals who are from or ancestrally descendent from Spanish-speaking countries (including Spain, excluding Brazil), while Laitno/a/x/e refers to individuals who are from or ancestrally descendent from the geographic region of Latin American (including Brazil, excluding Spain).[10] We recognize both Hispanic and Latino as pan-ethnic identities, as these terms reference a vast group of different cultural identities. Most individuals within those groups have more specific nuanced ethnic identities such as “Colombiano/a,” “Peruvian,” “Chicano/a,” “Nuyorican,” or “Tejano.” Hispanic or Latino/a/x/e individuals can be of any racial identity. For example, individuals who identify as “Black and Brazilian,” “White and Argentinian,” or “Asian and Guyanan” would all fall under the umbrella of Latino/a/x/e.[11]

**Appendix 2. Data extraction and analysis**

Section is adapted from: Martinez et a. "Conceptualization, operationalization, and utilization of race and ethnicity in major epidemiology journals 1995-2018: a systematic review." (2022) Am J Epidemiol. *In Press*.

Basic article characteristics were collected via PubMed and included title, first author, publication year, PMID, and journal. Additional information on study design (cohort, randomized control trial [RCT], ecological, case-control), data source, sample size, and health outcome type (health behavior, mental or physical health, healthcare access or utilization, other) were also collected. Specific article information related to each of the study's five research questions was then collected.

*Question 1: Inclusion of racial and ethnic data*. Reviewers were asked three yes/no questions for each article: "Did [the authors] measure RACE?", "Did [the authors] measure ETHNICITY?", "Did [the authors] combine RACE and ETHNICITY?". Reviewers were trained to select "yes" for the measurement of race if data on participant race was included in any capacity (e.g., in text or tables). The same instructions were given for ethnicity. Studies which treated Hispanic, Latino/x, African American or other ethnic identities as racial identities were recorded as "combining race and ethnicity". The same was true for articles which used "race" and "ethnicity" interchangeably.

*Question 2: Conceptualization of race and ethnicity*. For each article reviewers were asked to select yes/no for the question: "Did the authors provide a working definition of race?". To answer, reviewers looked for explicit definitions of race or statements which clearly indicated the author's perspective such as "race is a social construct…" or "biological traits such as race…". Definitions were recorded verbatim. The same questions were asked for ethnicity.

*Question 3: Operationalization*. Operationalization consists of both measurement and coding. For measurement, we employed Roth's (2016) framework which posits that race is a social construct that is fluid, multidimensional and can be broken down into numerous measures which each capture unique information about an individual's complex racial identity. Roth's six dimensions of race include identity, self-classification, observed, phenotype, reflected, and ancestry. Details of these dimensions are included in supplemental table 1. In addition to these six dimensions, reviewers for this study could also report that an article's measurement of race was "unclear/not stated" or "identity vs. self-classification" (unclear between identity and self-classification). We extrapolated Roth's framework and applied it to measurement of ethnicity. All applicable measures were selected for each article.

For coding, reviewers recorded the verbatim coding schemas reported by each article via free-response text boxes. If an article was previously recorded as combining race and ethnicity, then the same coding scheme was entered for both race and ethnicity. No attempt was made to collapse coding schemes and capitalization was not recorded.

*Question 4: Use in analyses*. Reviewers were asked to classify each article’s use of race and ethnicity in analyses as: “of interest”, “not of interest”, “exclusion”, or “other”. When race or ethnicity was used as a focal variable, for example in group comparisons, effect measure modification, mediation, or as an instrumental variable, “of interest” was selected. If race or ethnicity was as a matching criterion, confounder, or descriptive covariate, “not of interest” reviewers selected “not of interest”. “Exclusion” was selected if race or ethnicity was used to exclude participants from a study. The reference category of the racial, ethnic, or ethno-racial variable was recorded if race or ethnicity was included in a regression model.

*Question 5: Justification*. Reviewers were first asked “Did the authors provide a justification for any of their choices regarding race?” (yes/no). If “yes,” reviewers were instructed to copy-paste all justification from the text into a free-response text box. Reviewers were also asked “Did the authors justify their dimension (i.e., measure) of race?” (yes/no). A parallel set of questions were asked for ethnicity.

Data collection occurred between July 2019 and November 2021.

**Appendix Table 1. Definitions of race and ethnicity dimensions**

| | |
|---|---|
| **Dimensions of race*** | |
| Open-ended | Subjective self-identification assessed through an open-ended question (i.e., free response) |
| Close-ended | Self-identification assessed through a closed-ended question |
| Observed | Race identified by a third-party (e.g., interviewer) based on appearance alone or through interaction |
| Phenotype | Skin tone or other physical characteristics (e.g., hair texture, bone structure) assessed alone or in combination |
| Reflected | The race you believe others assume you to be; respondent's understanding of how they are view by others |
| Ancestry | As informed by familial history, genetic testing, or blood quantum |
| Unclear/not stated† | Insufficient or no information on how race was measured |
| Open vs close-ended† | Race was self-identified, but unclear if the question was open- or closed-ended |
| **Dimensions of ethnicity**** | |
| Open-ended | Subjective self-identification assessed through an open-ended question (i.e., free response) |
| Close-ended | Self-identification assessed through a closed-ended question |
| Country of origin | The country from which a person originally comes or “nationality” |
| Observed | Ethnicity identified by a third-party (e.g., interviewer) based on appearance alone or through interaction |
| Reflected | The ethnicity you believe others assume you to be; respondent's understanding of how they are view by others |
| Ancestry | As informed by familial history |
| Unclear/not stated† | Insufficient or no information on how ethnicity was measured |
| Open vs close-ended† | Ethnicity was self-identified, but unclear if the question was open- or closed-ended |

*Unless otherwise noted, dimensions originated from Roth (2016). **Ethnicity dimensions were adapted from Roth (2016); other dimensions of ethnicity may exist. †These dimensions were created by the authors for data collection purposes.

**Appendix Table 2. Characteristics of studies without race data *(N=94)***

| | **1995-99** | **2000-04** | **2005-09** | **2010-14** | **2015-18** |
|---|---|---|---|---|---|
| **No. Included** | 25 | 29 | 14 | 13 | 13 |
| **Study Design, No. (%)** | | | | | |
| RCT | 4 (16) | 5 (17) | 6 (43) | 3 (23) | 2 (15) |
| Cohort | 21 (84) | 21 (72) | 8 (57) | 10 (77) | 10 (77) |
| Case-control | 0 (0) | 1 (3) | 0 (0) | 0 (0) | 1 (8) |
| Ecological | 0 (0) | 2 (7) | 0 (0) | 0 (0) | 0 (0) |
| **Study Outcome, No (%)*** | | | | | |
| Health behavior | 1 (4) | 0 (0) | 0 (0) | 0 (0) | 1 (8) |
| Physical or mental | 20 (80) | 24 (83) | 10 (71) | 9 (69) | 8 (62) |
| Healthcare access | 1 (4) | 1 (3) | 2 (14) | 0 (0) | 1 (8) |
| Other | 3 (12) | 7 (24) | 3 (21) | 4 (31) | 3 (23) |
| **Sample size, No. (%)**** | | | | | |
| <1000 | 14 (54) | 18 (62) | 7 (32) | 1 (8) | 4 (31) |
| 1000-5000 | 6 (23) | 4 (14) | 3 (14) | 2 (15) | 2 (15) |
| 5001-10,000 | 1 (4) | 1 (3) | 1 (5) | 1 (8) | 1 (8) |
| 10,001-100,000 | 2 (8) | 3 (10) | 2 (9) | 5 (39) | 2 (15) |
| >100,000 | 1 (4) | 0 (0) | 9 (41) | 2 (15) | 4 (31) |
| Missing | 2 (8) | 3 (10) | 0 (0) | 2 (15) | 0 (0) |

*Study outcomes were classified as health behaviors (e.g., smoking, dietary intake, physical activity, sexual behaviors), mental or physical health (e.g., obesity, high blood pressure, cancer, depression), health care access of utilization (e.g., health insurance status, number of primary care visits, quality of care), or other. Study outcomes are not mutually exclusive and may sum to more than 100%. **Some studies listed more than one analytic sample size; values may sum to more than 100%.

**Appendix Table 3. Racial coding schemes (N=62)**

| Coding Scheme | No. | (%) |
|---|---|---|
| black, other, white | 9 | (15) |
| black, white | 9 | (15) |
| non-white, white | 6 | (10) |
| nonwhite, white | 5 | (8) |
| ns | 5 | (8) |
| white | 5 | (8) |
| asian, black, other, white | 4 | (6) |
| black | 3 | (5) |
| asian, black, mixed race or missing data, native american, white | 2 | (3) |
| other, white | 2 | (3) |
| american indian or alaskan native, asian or pacific islander, black, white | 1 | (2) |
| american indian, asian, black, white or other | 1 | (2) |
| black race, non-black race | 1 | (2) |
| black, mixed/other, not reported, white | 1 | (2) |
| black, nonblack | 1 | (2) |
| black, not stated, other, white | 1 | (2) |
| black, of asian descent, white | 1 | (2) |
| black, other minority, white | 1 | (2) |
| black, other race, white | 1 | (2) |
| black, other | 1 | (2) |
| blacks, non-blacks | 1 | (2) |
| japanese, other | 1 | (2) |

Across stratum, 22 unique racial coding schemes were identified from amongst 62 coding schemes belonging to 62 studies. These studies included racial data and may have included ethnicity data, but did not combine the two into an ethno-racial construct. Capitalization was not collected. No attempt was made to collapse coding schemes based on similarity.

### Appendix Table 4. Ethnoracial coding schemes (N=86)

| Coding Scheme | No. | (%) |
|---|---|---|
| black, hispanic, other, white | 9 | (14) |
| asian, black, hispanic, other, white | 5 | (8) |
| black, hispanic, white | 4 | (6) |
| black, other, white* | 3 | (5) |
| american indian, asian/pacific islander, black, hispanic, unknown, white | 2 | (3) |
| asian or pacific islander, black, hispanic, other, white | 2 | (3) |
| hispanic, non-hispanic black, non-hispanic white, other | 2 | (3) |
| mexican american, non-hispanic black, non-hispanic white, other | 2 | (3) |
| non-white, white* | 2 | (3) |
| ns | 2 | (3) |
| african american, american indian, asian or pacific islander, hispanic, white | 1 | (2) |
| african american, american indian, asian, white | 1 | (2) |
| african american, american indian, white | 1 | (2) |
| african american, asian, hispanic, non-hispanic white, other (multiracial, american indian/alaskan native, and native hawaiian/other pacific islander) | 1 | (2) |
| african american, asian, hispanic, non-hispanic, other, white | 1 | (2) |
| african american, asian, hispanic, other, white | 1 | (2) |
| african american, asian, latina, other, white | 1 | (2) |
| african american, asian, other/multiracial, white | 1 | (2) |
| african american, hispanic, other (asian, other races), white | 1 | (2) |
| african american, hispanic/other, white | 1 | (2) |
| african american, japanese american, latino, native hawaiian, white | 1 | (2) |
| african american, non-hispanic white | 1 | (2) |
| african american, non-hispanic white or mexican american | 1 | (2) |
| african american, other, unknown, white | 1 | (2) |
| african-american, asian, hispanic, other, white | 1 | (2) |
| african-american, mexican-american | 1 | (2) |
| african-american, mexican-american, white | 1 | (2) |
| aleut, asian, black, 'don't know or no answer', hispanic, white | 1 | (2) |
| am indian or alaska native, asian, black, hispanic or latino, multiple races, native hawaiian or other pacific islander, white | 1 | (2) |
| american indian, asian, black, hispanic, missing, white | 1 | (2) |
| american indian/alaska native, asian american, black, hispanic/latino, non-hispanic white | 1 | (2) |
| american indian/alaska native, asian/pacific islander, black, hispanic/latino, native hawaiian, not-reported, white non-hispanic | 1 | (2) |
| ashkenazi jew, black, other, white | 1 | (2) |
| asian or pacific islander, declined response, hispanic, multiethnic, native american, non-hispanic black, non-hispanic white | 1 | (2) |
| asian, black, hispanic, native american, other, pacific islander, white | 1 | (2) |
| asian, black, hispanic, native american, other, white | 1 | (2) |
| asian, black, hispanic, other (native american, other, unknown), white | 1 | (2) |
| asian, black, hispanic, white | 1 | (2) |
| asian, black, other non-underrepresented minority, other underrepresented minority, white | 1 | (2) |
| asian, black, white, white with hispanic ethnicity | 1 | (2) |
| asian, hispanic, non-hispanic black, non-hispanic white | 1 | (2) |

| | | |
|---|---|---|
| asian, hispanic, non-hispanic black, non-hispanic white, other | 1 | (2) |
| asian/pacific islander, black, hispanic, other, white | 1 | (2) |
| asian/pacific islander/native american, hispanic, non-hispanic black, non-hispanic white | 1 | (2) |
| black ethnic origin | 1 | (2) |
| black non- hispanic, hispanic, other, white non-hispanic | 1 | (2) |
| black non-latino, latino, other, white non-latino | 1 | (2) |
| black or african american, hispanic ethnic origin, other, white | 1 | (2) |
| black or african american, hispanic or latino, not hispanic or latino, white | 1 | (2) |
| black, hispanic and other, non-hispanic white | 1 | (2) |
| black, hispanic, mixed, other, white | 1 | (2) |
| black, latina, other, white non latina | 1 | (2) |
| black, latino/hispanic, other, white | 1 | (2) |
| black, other, white non-hispanic* | 1 | (2) |
| ethnic/racial minority, white | 1 | (2) |
| hispanic, missing data, non-hispanic black, non-hispanic white, other | 1 | (2) |
| hispanic, non-hispanic asian or pacific islander, non-hispanic black, non-hispanic white | 1 | (2) |
| hispanic, non-hispanic asian, non-hispanic black, non-hispanic white | 1 | (2) |
| non-hispanic white, not non-hispanic white | 1 | (2) |
| non-hispanic white, other* | 1 | (2) |
| non-hispanic whites, ns* | 1 | (2) |
| nonwhite, white* | 1 | (2) |
| other, white* | 1 | (2) |

Across stratum, 63 unique racial coding schemes were identified from amongst 86 total coding schemes belonging to 86 studies. These 86 studies combined the racial and ethnic data into an ethno-racial construct or operationalized an ethno-racial construct. (*) These coding schemes appear identical to a few the racial coding schemes (Supp. Table 3); studies associated with these coding schemes did use an ethno-racial construct but the variable recoding obscured this. For example, a study may treat "Hispanic" or "Latino" as a racial category and then recode to a binary variable of "non-white, white," where "non-white" includes "Hispanic, Black, Asian, and Native American/Alaskan Native" individuals. Capitalization was not collected. No attempt was made to collapse coding schemes based on similarity.

**Appendix Table 5. Role of race and/or ethnicity in analyses, 1995-2018**

| | 1995-99 | 2000-04 | 2005-09 | 2010-14 | 2015-18 |
|---|---|---|---|---|---|
| **No. Included** | 20 | 31 | 28 | 33 | 37 |
| **Analytic role, No. (%)** | | | | | |
| "Of interest" | 2 (10) | 4 (13) | 7 (25) | 5 (15) | 5 (14) |
| "Not of interest" | 17 (85) | 26 (84) | 18 (64) | 28 (85) | 31 (84) |
| "Exclusion" | 1 (5) | 0 (0) | 2 (7) | 0 (0) | 1 (3) |
| "Other" | 0 (0) | 1 (3) | 1 (4) | 0 (0) | 0 (0) |

No. Included indicates the number of articles which included data on race and/or ethnicity.

### Appendix 3. Secondary analysis: JAMA publications

**Table 1. Characteristics of JAMA articles (*N=113*)**

| | 1995-99 | 2000-04 | 2005-09 | 2010-14 | 2015-18 |
|---|---|---|---|---|---|
| **No. Included** | 25 | 33 | 18 | 20 | 17 |
| **Studies including, No. (%)** | | | | | |
| Race data | 11 (44) | 19 (58) | 12 (67) | 15 (75) | 13 (76) |
| Ethnicity data | 5 (20) | 9 (27) | 10 (56) | 12 (60) | 11 (65) |
| Both race & ethnicity data | 5 (20) | 9 (27) | 10 (56) | 12 (60) | 11 (65) |
| **At least one justification, No. (%)** | 3 (27) | 2 (11) | 8 (67) | 7 (47) | 5 (38) |

**Table 2. Measures of race and ethnicity over time, 1995-2018**

| | 1995-99 | 2000-04 | 2005-09 | 2010-14 | 2015-18 |
|---|---|---|---|---|---|
| **No. Included race data** | 11 | 19 | 12 | 15 | 13 |
| **Measure of race, No. (%)** | | | | | |
| Open-ended, self report | 0 (0) | 0 (0) | 0 (0) | 0 (0) | 1 (8) |
| Close-ended, self report | 0 (0) | 2 (11) | 1 (8) | 1 (7) | 6 (46) |
| Observed | 0 (0) | 0 (0) | 1 (8) | 0 (0) | 0 (0) |
| Phenotype | 0 (0) | 0 (0) | 0 (0) | 0 (0) | 0 (0) |
| Reflected | 0 (0) | 0 (0) | 0 (0) | 0 (0) | 0 (0) |
| Ancestry | 0 (0) | 0 (0) | 0 (0) | 0 (0) | 0 (0) |
| Unclear/not stated | 11 (100) | 17 (89) | 7 (58) | 9 (60) | 5 (38) |
| Open vs close-ended* | 0 (0) | 0 (0) | 3 (25) | 5 (33) | 1 (8) |
| **No. Included ethnicity data** | 5 | 9 | 10 | 12 | 11 |
| **Measure of ethnicity, No. (%)** | 0 (0) | 0 (0) | 0 (0) | 0 (0) | 0 (0) |
| Open-ended, self-report | 0 (0) | 0 (0) | 0 (0) | 0 (0) | 1 (9) |
| Close-ended, self-report | 0 (0) | 2 (22) | 2 (20) | 1 (8) | 6 (55) |
| Country of origin | 0 (0) | 0 (0) | 0 (0) | 0 (0) | 0 (0) |
| Observed | 0 (0) | 0 (0) | 0 (0) | 0 (0) | 0 (0) |
| Reflected | 0 (0) | 0 (0) | 0 (0) | 0 (0) | 0 (0) |
| Ancestry | 0 (0) | 0 (0) | 0 (0) | 0 (0) | 0 (0) |
| Unclear/not stated | 5 (100) | 7 (78) | 5 (50) | 9 (75) | 3 (27) |
| Open vs close-ended* | 0 (0) | 0 (0) | 3 (30) | 2 (17) | 1 (9) |

Selections of multiple measures was allowed; percents may sum to >100%. *For racial "open vs close-ended" race was noted as self-reported by a participant, but it was unclear if the question was open- or closed-ended. Same applies to ethnic "open vs close-ended."
